# Developing Cardiac Digital Twins at Scale: Insights from Personalised Myocardial Conduction Velocity

**DOI:** 10.1101/2023.12.05.23299435

**Authors:** Shuang Qian, Devran Ugurlu, Elliot Fairweather, Marina Strocchi, Laura Dal Toso, Yu Deng, Gernot Plank, Edward Vigmond, Reza Razavi, Alistair Young, Pablo Lamata, Martin Bishop, Steven Niederer

## Abstract

Large-cohort studies using cardiovascular imaging and diagnostic datasets have assessed cardiac anatomy, function, and outcomes, but typically do not reveal underlying biological mechanisms. Cardiac digital twins (CDTs) provide personalized physics- and physiology-constrained *in-silico* representations, enabling inference of multi-scale properties tied to these mechanisms.

We constructed 3464 anatomically-accurate CDTs using cardiac magnetic resonance images from UK biobank and personalised their myocardial conduction velocities (CVs) from electrocardiograms (ECG), through an automated framework.

We found well-known sex-specific differences in QRS duration were fully explained by myocardial anatomy, as CV remained consistent across sexes. Conversely, significant associations of CV with ageing and increased BMI suggest myocardial tissue remodelling. Novel associations were observed with left ventricular ejection fraction and mental-health phenotypes, through a phenome-wide association study, and CV was also linked with adverse clinical outcomes.

Our study highlights the utility of population-based CDTs in assessing intersubject variability and uncovering strong links with mental health.

## Introduction

Large cohort multi-modality cardiovascular imaging and diagnostic data sets are increasingly available and are being used to link heart anatomy and function with physiological, lifestyle, and clinical outcomes^1–3^. While providing hypothesis-generating correlations, they have been less successful at identifying the underlying mechanisms that drive these correlations. This is in part because they are restricted to the analysis of observed attributes and do not identify the underlying physiology that may explain the observed correlations.

A strategy to alleviate this limitation is the personalization of Cardiac Digital Twins (CDTs)^4–6^. CDTs provide physics- and physiology-constrained *in-silico* representations of specific individuals. They are personalized by integrating multimodal data, enabling multi-scale structure and function to be inferred from clinical measurements. The model parameters that explain the data become the attributes that describe the underlying physiology. Early forms of CDTs have shown great potential in supporting clinical decision-making and providing tailored therapies as in prospective studies of ventricular tachycardia^7^, atrial fibrillation^8^ and cardiomyopathy^9^.

However, creation of CDTs is associated with challenges. Complex data processing and specialist methodology and requirement of large computational resources are required, which limit their broad adoption in both industrial and clinical settings. Presently, studies are constrained to working with small patient cohorts with at most 100 patients^10^, limiting their application and scalability to large population datasets.

The creation of CDTs involves two crucial steps. First is the construction of the anatomical twin, the computational replica of the anatomical structures of each subject’s heart from medical images. Previously, semi-automatic workflows of heart mesh generation have been developed^11,12^, demanding substantial computational resources and considerable manual interventions by trained experts. The second is to build the functional twin, i.e. identifying bespoke electrophysiological parameters that replicate clinical measurements, for example, electrocardiograms (ECGs). This step is often more challenging, requiring numerous computationally intensive simulations to calibrate the multi-scale parameters, based on the specific biophysical fidelity needed. Parameter personalization remains an ongoing challenge, with the majority of modelling studies relying on ‘average’ parameters derived from the literature^7,13,14^.

The feasibility of generating CDTs at scale hinges on the development of a computationally and time-efficient automated workflow for both CDT creation steps. Recent advances in image segmentation, including nnU-Net^15^ and Atlas-based approaches^16^ provide robust and precise 3D anatomical structures within abbreviated timeframes, facilitating the rapid creation of anatomical meshes from medical images. The emergence of surrogate models as novel statistics and machine learning tools, for example, Gaussian Process Emulators (GPE)^17^, provide a low-cost statistical representation of computationally expensive models. Such surrogate models enable global sensitivity analysis to identify important model parameters and therefore constrain the viable parameter space, which can reduce the number of simulations necessary for model calibration.

In this study, we developed a methodology that integrates multi-modality data from the UK biobank^18^ within a CDT framework and first demonstrated the feasibility of CDT creation at scale. Within this framework, we inferred a key myocardial tissue electrical property, myocardial conduction velocity (CV), for each CDT. We then reported how CV varies across sex, body mass index and age, together with imaging and ECG-derived phenotypes (biventricular myocardial mass and QRS duration (QRSd) respectively). We also conducted a phenome-wide association study to explore their relationships with other phenotypes reported in the UK Biobank, followed by assessing their ability to independently predict clinical outcomes.

## Results

### Anatomical and functional CDT generation workflow

The CDTs were built from UK Biobank magnetic resonance imaging (MRI) and ECG data sets as depicted in Fig. 1. Each CDT replicates the ECG QRSd and consists of a model of the heart anatomy, the preferred myocyte orientation or fibre structure, a fast endocardial conducting layer, the location of the activating Purkinje fascicles, the activation timing, the tissue conductivity, the degree of anisotropy, and the location of the virtual ECG electrodes. The personalization involved the generation of the bespoke bi-ventricular anatomy from MRI and the inference of the CV that replicates the ECG QRSd. The anatomical personalization of a CDT, the first step in the pipeline, took 5 minutes to be built from MRI images (avg mesh resolution 0.9mm) on a desktop with 16 cores.

**Fig. 1:**
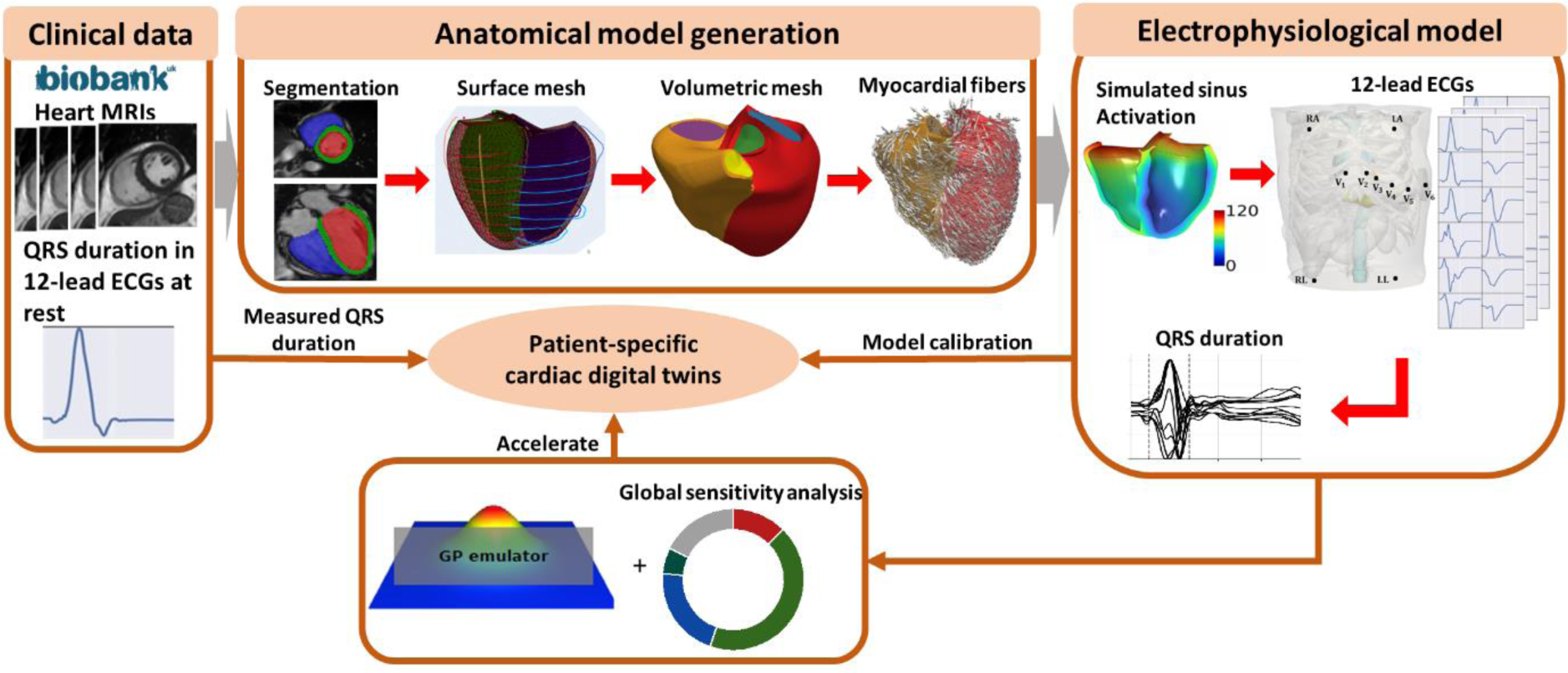
The automated anatomical and functional (Electrophysiological) CDT generation workflow. The anatomical models are personalised finite element meshes with physiological-detailed myocardial fibres constructed from the short-axis and long-axis heart images in the UK biobank following the steps of segmentations, surface meshes and volumetric meshes construction as well as myocardial fibre generation. The functional CDT workflow is to replicate the electrophysiological activities within the anatomical models to match the QRS duration from the clinically measured 12-lead ECGs.

The functional personalization required a preliminary analysis to determine which parameters could be inferred from the available data and which needed to be set to reference prior values. Thus, we performed two global sensitivity analyses. In 10 representative cases sampled across sex, BMI, and age, we calculated the sensitivity of the QRSd prediction to 20 tissue-level physiological parameters (Fig. 2a and Supplementary Table 2) or 30 ECG electrode positions combined with the most important tissue-level property (Fig. 2b; Supplementary table 3). Notably, the first analysis revealed that CV has the dominant effect on QRSd, accounting for 72.7 ± 3.2 % the sensitivity, across all electrophysiological parameters, see Fig. 2. The second analysis then studied the potential impact of ECG electrode positions, finding that CV affects the QRSd the most (67.2 ± 10.4 %).

**Fig. 2:**
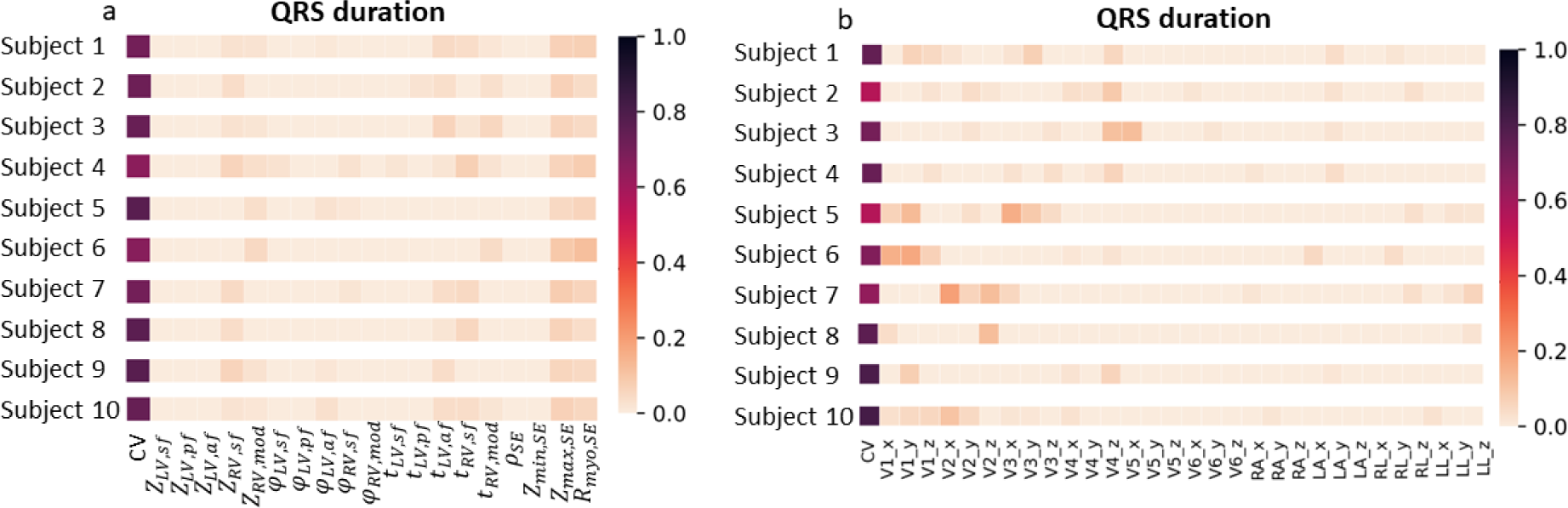
Global sensitivity analysis. The total effects of input EP parameters explain the variance of output QRS duration in 10 subjects sampled from the cohort based on sex, age and BMI. a is for only tissue-level EP parameters and b is for the 30 ECG electrodes’ location parameters combining with the most important tissue-level parameter from a.

Given CV was identified as the only key parameter that explains most of the variation in QRSd, we chose to calibrate the models by applying a computational-efficient bisection method to search for the personalized CV (constrained by physiological measurements in literature ^19,20^) to match with measured QRSd from the 12-lead ECGs at rest during sinus rhythm. The CDT calibration process assumed all other parameters were set to reference prior values taken from the literature, introducing an estimated uncertainty of -13% to +15.6% around the ‘true’ CV (Supplementary Figure 2). The personalised CV parameter was inferred in 8 minutes on a desktop with 16 cores.

We used the 4,329 first participants from the UK biobank that had adequate geometrical information^16^ and reported QRSd, sex, age, BMI/weight and height information. Of these, 3945 (91.1%) and 3464 (80%) were successfully processed through the anatomical and functional personalization workflows. Summary participant characteristics are shown in Supplementary Table 1.

### Model validation

To validate the CDT workflow, we compared the QRS morphology in simulated ECGs against the measured ECGs in the 10 representative subjects. As the ECGs were simulated from a reference torso and heart location, we do not expect perfect matches in all leads. We have adopted a lead-to-lead comparison approach used to compare ECGs clinically^21^. To quantify the ability of the model to replicate the ECG morphology, we plotted the simulated 12 lead ECGs (filtered, scaled, and temporally aligned) against measured ECG and correlation coefficients (*r*) were calculated. Supplementary Figure 3 shows the comparison, with plots ordered in descending order of the correlation coefficients. 56% of ECG leads in all subjects matched with the recordings well (*r* > 0.5). Supplementary Figure 4 shows the averaged *r* of the best correlated ECG leads as the number of leads considered increases. The maximum average *r* considering only the best correlated ECG leads was 0.95, while its value dropped to 0.79 when considering the top 6 ECG leads.

To further validate our modelling approach, we investigated whether subjects with pathologically slow conduction, for instance, caused by fascicular block or heart failure, can be differentiated through their personalized CVs. Those subjects were identified using the summary diagnoses for hospital inpatients in the UK Biobank with the specific disease types as shown in Supplementary Table 5. Fig. 3 shows that subjects with fascicular block (N=46) had 16.8% lower CV (0.482± 0.11 vs 0.579±0.08 m/s, *P* = 1.1 × 10^−14^), and 23.3% longer QRSd (108.4±26.0 vs 87.9±12.6ms, *P* = 2.2 × 10^−26^), comparing to their counterparts. Similarly, we also observed 5.2% slower CV (0.548± 0.10 vs 0.578±0.08m/s, *P* = 6.6 × 10^−3^) and 10.7% longer QRSd (97.6±20.8 vs 88.1±12.8ms, *P* = 2.1 × 10^−8^) in subjects with heart failure (N=60), comparing to their counterparts.

**Fig. 3:**
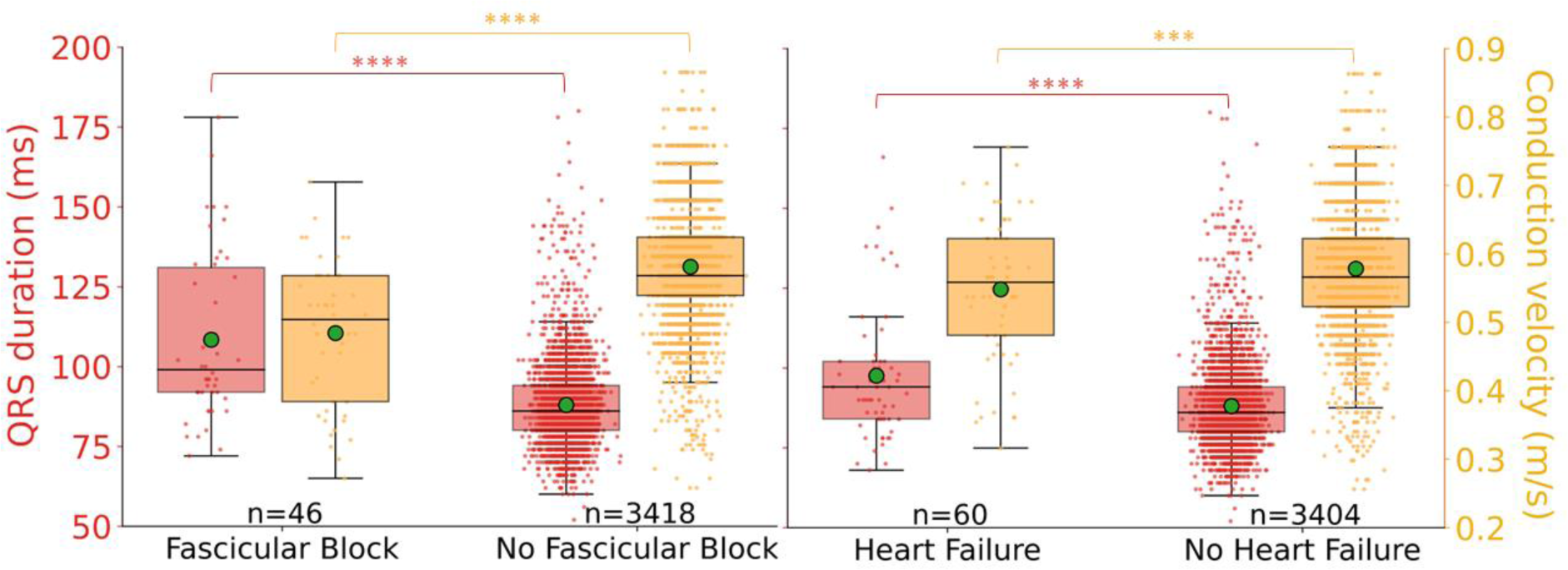
Boxplots of QRS duration (red) and conduction velocity (yellow) for comparing groups of participants afflicted with fascicular block and non-afflicted counterparts. The green dots indicate the mean. The corresponding P values are from the student’s t-test. *P<0.05; **P<0.01; ***P<0.001; ****P<0.0001.

### Comparison for different sex, BMI and age groups

We compared QRSd, CV and biventricular myocardial mass, categorized into different groups of sex, BMI and age as shown in Fig. 4. The QRSd was 9.4% longer in males (92.9±13.2 vs 84.2±11.5 ms, *P* = 2.5 × 10^−89^). This is consistent with males tending to have larger hearts (male: 180.9±28.5g vs 131.1±18.9g, *P* = 0). However, CVs were the same in men and women (0.576±0.09 vs 0.579±0.08m/s, P=0.25).

**Fig. 4:**
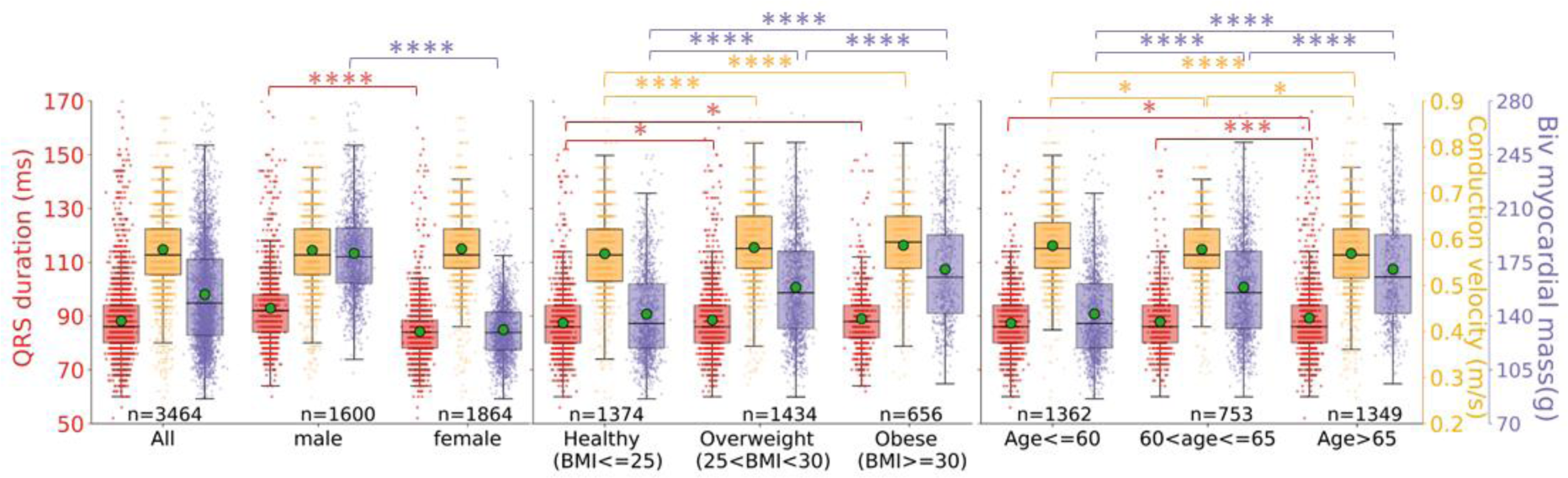
Boxplots of QRS duration (red), conduction velocity (yellow) and myocardial mass(purple) for different groups of sex, BMI and age. The green dots indicate the mean. The corresponding P values are from the student’s t-test. *P<0.05; **P<0.01; ***P<0.001; ****P<0.0001.

QRSd was longer for overweight and obese (25 ≤ *BMI* ≤ 30 and *BMI* ≥ 30) compared to healthy (*BMI* ≤ 25) groups (overweight: 88.6±12.9 and obese: 89.1±13.0, vs healthy: 87.5±13.2ms, P=0.03 and 0.01). Again, this increase in QRSd with BMI was reflected with a corresponding increase in heart size (healthy: 141.4±29.2, overweight: 158.7±33.5, obese: 170.5±36.9g, all *P* < 1 × 10^−13^). In contrast with sex, this increase in QRSd was also associated with a progressive increase in CVs (healthy: 0.569±0.08m/s vs overweight and obese: 0.582±0.08 and 0.587±0.09m/s, *P* = 1.8 × 10^−5^ and 7 × 10^−6^), which suggests the compensatory mechanism for the increment of heart size (i.e. CV is increased as a mechanism to reduce QRSd when the heart needs to grow to meet the larger demand of an increased BMI).

QRSd increased with ageing, which was significant when comparing the *Age* ≤ 60 group (87.3±11.6ms) with *Age* ≥ 65 (87.9±12.0ms, P=0.03) and 60 ≤ *Age* ≤ 65 (89.3±14.8ms, *P* = 1.1 × 10^−4^). This QRS increase is consistent with an observed CV decrease (*Age* ≤ 60: 0.586±0.08 vs 60 ≤ *Age* ≤ 65: 0.578±0.08 vs *Age* ≥ 65: 0.569±0.09m/s, all P<0.02), and myocardial mass increase (141.4±29.2 vs 158.7±33.5 vs 170.5±36.9g, all *P* < 1 × 10^−13^). By inferring CV, it is possible to determine when changes in QRSd with age, sex or BMI are caused by changes in cardiac anatomy or biological material properties.

### Phenome-wide association study

We performed a phenome-wide association study (PheWAS) to explore the correlations between the multimodal and CDT-derived phenotypes and 473 UKBB reported phenotypes in categories: pulse wave analysis, LV size & function (automatically derived from heart MRIs), abdominal composition, medication (medical treatment received), primary demographics, early-life information, self-reported medical conditions, lifestyle diet, alcohol, smoking, physical activity, physical measures, education and employment, mental health, and clinical outcomes of seven common diseases categorized from summary diagnoses in UKBB (Supplementary Table 5).

Fig. 5 shows the Manhattan plot of the univariate correlation P values (two-sided) between M = 3 multimodal phenotypes (myocardial mass, QRSd and inferred CV) and N = 473 UKBB phenotypes for M × N = 1419 times, with 87 correlations reaching the Bonferroni threshold for multiple comparisons (*P_bonf_* = 3.5 × 10^−5^ for α = 0.05) and 146 correlations reaching the false-discovery rate (FDR) threshold (*P_fdr_* = 0.005 for α = 0.05). For the correlation coefficients in the Manhattan plot see Supplementary Figure 6.

**Fig. 5:**
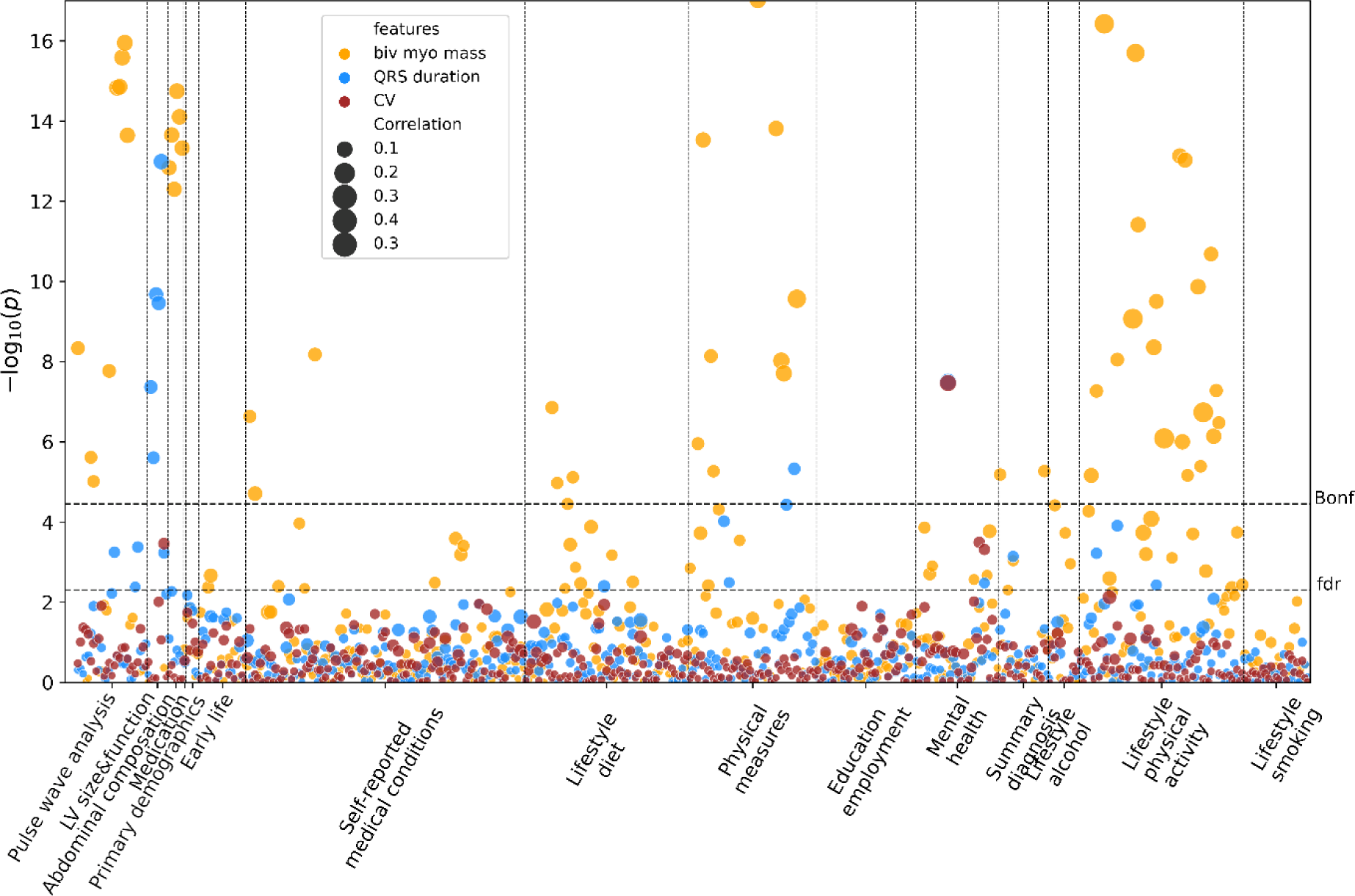
Manhattan plot showing the −*log*_10_*P* (two-sided t-test) for correlations between the QRS duration, CV and myocardial mass, and UKBB reported phenotypes. The size of the dots indicates the absolute Pearson’s correlation coefficient. The dashed horizontal lines are the Bonferroni threshold (Bonf) and the false-discovery rate (fdr) (α = 0.05). Note the plot is clipped at 17 for better visualization for QRS duration and CV. The complete plot is in the Supplementary Figure 5.

The QRSd was significantly associated with seven cardiac structural and functional phenotypes including LV end-diastolic, end-systolic and stoke volumes, cardiac output, pulse/heart rate (−*log*10*P* ≥ 2.4, |*r*| ≥ 0.05) as well as LV ejection fraction (−*log*10*P* = 3.2, r=-0.06). In contrast, CV was only significantly associated with LV ejection fraction (−*log*_10_*P* = 3.5, r=0.06). The biventricular myocardial mass was significantly associated with all phenotypes mentioned above (−*log*_10_*P* > 9.6, |*r*| ≥ 0.16) where higher correlations were seen for structural phenotypes such as LV stroke, end diastolic and end-systolic volumes (−*log*10*P* > 68.8, |*r*| > 0.3).

Interestingly, the three phenotypes were all significantly associated with mental health-related phenotypes. Both QRSd and CV were significantly associated with ‘the longest period of depression’ (both: −*log*_10_*P* = 7.5; QRSd: r=0.15, CV: r=-0.15) and ‘Seen doctor (GP) for nerves, anxiety, tension or depression’ (QRSd:−*log*_10_*P* = 2.5, r=0.05; CV: −*log*_10_*P* = 3.3, r=-0.06). CV and myocardial mass were significantly associated with ‘Seen a psychiatrist for nerves, anxiety, tension or depression’ (CV: −*log*10*P* = 3.5, r=-0.06; myocardial mass: −*log*10*P* = 2.6, r=-0.05). Consistent with previous studies^1^, we find that myocardial mass was solely associated with the Neuroticism score, Frequency of depressed mood in the last 2 weeks, and Happiness (2.6 < −*log*_10_*P* < 3.9, r: [-0.1, -0.05]).

### Association with clinical diagnoses

We investigated the associations of the multimodal and CDT-derived phenotypes with seven common diseases, categorized using the summary diagnoses for hospital inpatients. We trained a logistic regression model on QRSd, myocardial mass and CDT-derived CV to predict disease occurrence, adjusted with demographics/anthropometrics factors including sex, age BMI, age*BMI and sex*age. Fig. 6 and Supplementary Table 6 presents the odd ratios (ORs) derived from the regression coefficients (represented as *value*_[95%*confidence interval*]_) and the corresponding P values.

**Fig. 6:**
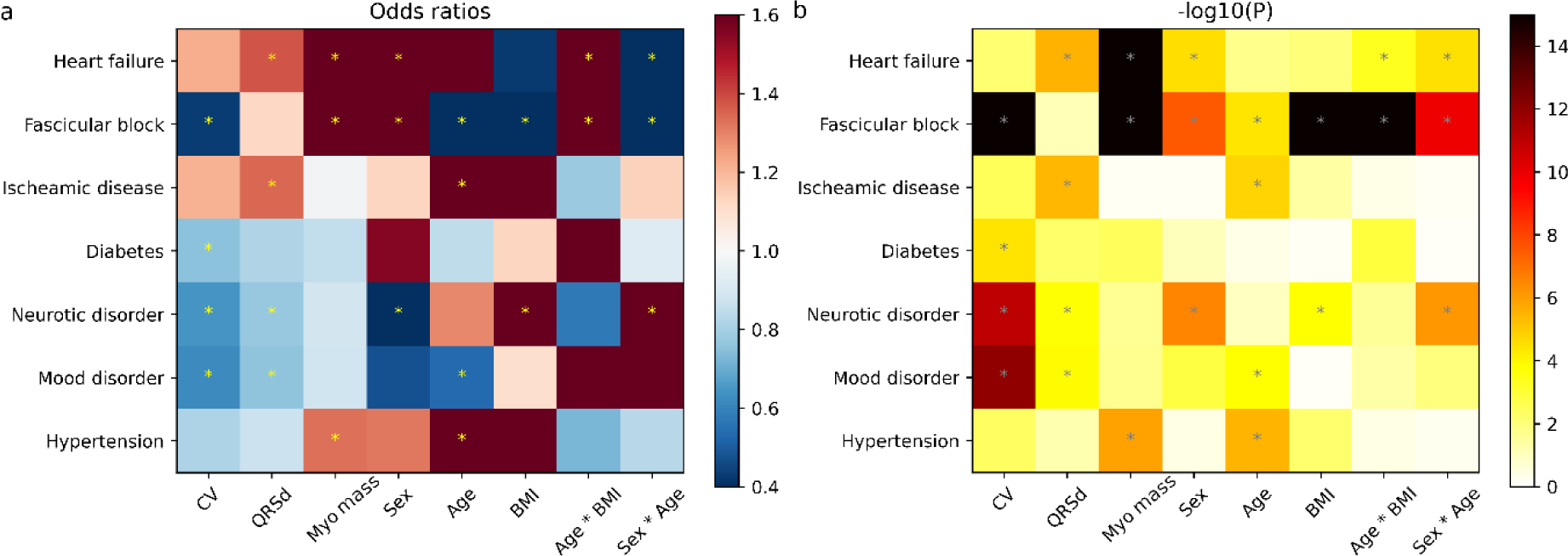
Association of CV, QRS duration and myocardial mass with common diseases. (1) Odd ratios for phenotypes as input risk factors for common diseases as the outcomes. Sex, age, BMI, age*BMI and sex*age were included in the logistic regression analysis (N=3464). (2) The corresponding P values (two-sided t-test) for odd ratios. *Result reaches Bonferroni threshold (*P*_*Bonf*_ = 6.9 × 10^−4^ for *a*=0.05).

Smaller CV and greater myocardial mass were associated with a high risk of Fascicular block (ORs=0.43_[0.37,0.51]_ and 1.72_[1.54,1.92]_, both *P* < 1 × 10^−16^). In contrast, both greater QRSd and myocardial mass were associated with a higher risk of heart failure (ORs=1.38_[1.20,1.58]_ and 1.66_[1.50,1.83]_, *P* = 3 × 10^−6^ *and P* < 1 × 10^−16^). Ischaemic diseases were more likely to develop with greater QRSd (ORs=1.34_[1.19,1.52]_, *P* = 4 × 10^−6^), while diabetes was more likely to develop with smaller CV (ORs=0.75_[0.66,0.86]_, *P* = 3.6 × 10^−5^). Reduced CV was a better predictor than QRSd for both neurotic and mood disorders, (CV: ORs=0.65_[0.57,0.73]_ and 0.62_[0.54,0.72]_, *P* < 1.07 × 10^−11^vs QRSd: ORs= 0.77_[0.68,0.88]_ and 0.76_[0.66,0.87]_ *P* < 1.27 × 10^−4^). The extended analysis (Supplementary Figure 7) shows CV was significantly lower in participants having mood disorders than their counterparts (0.564±0.085 vs 0.578± 0.085m/s, P=0.03), while QRSd and myocardial mass did not (QRSd: 88.8± 14.2 vs 88.2±13.0ms, P=0.57 and vol: 142.3± 30.5 vs 147± 32.9g, P=0.07). Hypertension was solely associated with greater myocardial mass (OR=1.33_[1.18,1.49]_, *P* = 1.26 × 10^−6^).

## Discussion

This study makes three major contributions. First, we have developed a fully automated CDT generation workflow including both patient-specific anatomical and electrophysiological models, using multimodal data including both medical imaging and clinically measured ECGs. This workflow is the first of its kind to construct biophysically detailed CDTs at a large scale, benefiting from using statistical and AI tools that allow global sensitivity analysis on all known model parameters.

Second, we have showcased the capacity of utilizing the quantitative CDT-derived phenotype to unveil the underlying biological mechanisms and elucidate the variability in observable attributes such as imaging and ECG phenotypes among different populations according to sex, age and BMI. Specifically, we show that well-known sex-specific differences in QRSd can be fully explained by myocardial anatomy and that CV is the same in men and women, and that myocardial remodeling leads to changes in CV associated with ageing and increased BMI.

Third, we performed a large-scale PheWAS study and found more significant associations of the CDT-derived phenotype with cardiac and mental-health phenotypes relating to neurological disorders, including depression, compared against known ECG and imaging phenotypes. Furthermore, we found a stronger association between the decreased CV and a higher risk of neurotic and mood disorders, compared to the myocardial mass and QRSd.

These results demonstrate the usefulness of generating CDT-derived phenotypes, even from the UK biobank where the majority of participants identified as healthy. The ability to identify the electrophysiological biomarkers: myocardial CV at scale could help identify potential therapeutic targets and evaluate the therapeutic potential (or side effects) of existing drugs and heart disease medications for mental health and neurodegenerative disorders.

CDTs are rapidly advancing in recent decades, leveraging their capacity to guide, inform, monitor, diagnose and prognose therapies and surgical interventions in many current prospective clinical studies^22^, paving the way for moving into industrial and clinical settings^4^. This shift requires a step change in the speed, robustness, validation, and uncertainty quantification in both anatomical and functional model creation workflow.

To date, a wide range of automated anatomical model creation workflows based on machine learning have been proposed allowing large-scale population analysis such as on the UK Biobank dataset^1,23,24^, however, these models only account for surfaces depicting the overall heart structure, but not high-quality volumetric meshes. There are also volumetric mesh creation workflows existing for atria^25,26^, ventricles^12^ and whole hearts^11,27^, but these have only been applied to smaller datasets (<100) due to the high computational costs and manual steps required. Here we reported a fully automated volumetric mesh generation workflow that can create personalized anatomical meshes at scale within clinical timescales (∼5 minutes/CDT).

Aside from anatomical model personalization, functional model personalization is more challenging, where most current modelling studies were developed using ‘average’ material property values from broader physiological studies with limited personalization^28,29^. Recent developments in high fidelity biological and physiological electrophysiological CDT frameworks incorporated detailed features like the His-Purkinje fascicles to replicate detailed QRS complex morphology ^12,30^. However, only a subset of parameters underwent personalization, yet still demanding significant computation resources and time. Alternative computationally efficient approaches based on machine learning and statistical methods were reported but often fall short in capturing all anatomical/functional details and struggle to generalize effectively^31,32^.

To move to population-level studies, it is of critical importance to balance biophysical fidelity, parameter inference and computational cost. In this work, instead of making CDTs to replicate recorded ECG morphologies that are more likely to be afflicted by subject-specific noises, we constructed feature-specific CDTs to replicate the QRSd which can be extracted from models robustly and computationally-efficiently^33^. Similar to previous biophysical-detailed frameworks^12^, our CDT EP framework incorporates parameters encapsulating knowledge derived from physiological and histological/anatomical experiments in the past. Our sensitivity analysis suggests that an optimal trade-off between model fidelity and parameter identifiability is to restrict the EP personalization to one single parameter, CV, showing a dominating effect on QRSd (>67.2%), ensuring its computational efficiency (∼8 mins/CDT).

To truly realize the predictive capability of developed CDTs, it is important to determine the bounds of fidelity and validity of our CDTs (i.e. which aspects of the real-world system are sufficiently recapitulated?). We have conducted two model validations. First, we compared how well our simulated 12-lead ECGs reproduce the recordings lead-wisely by conducting correlation tests on the 10 representative subjects. We found good correlations in 56% of lead ECGs (*r* > 0.5) from all subjects, as we expected. We have standardized the ECG electrode locations due to data unavailability, where such information has been previously shown to be the main driver of QRS complex morphological difference, but not for QRSd^29,34^. Therefore, we can conclude our CDT framework is still reliable with good accuracy for QRSd. Second, our CDT approach was tested to identify the CVs of subjects afflicted with fascicular block or heart failure. These are two conditions with known slow cardiac conduction substrates, and we found that their CV was indeed significantly slower than the counterparts by 16.8% and 5.2% (Fig. 3), given we used a unified healthy conduction system in our personalized QRSd calibration process. An additional positive validation result is the fact that the separate association test on clinical outcomes identified that lower CV is associated with a much higher risk of fascicular block (Fig. 6). Further, in a verification test, we quantified the uncertainty of the inferred CVs to be <15.6% variation around the ‘true’ value, considering the uncertainty in assuming all other parameters (electrophysiology and ECG electrodes positioning) as reference prior values, which further increases the credibility of our approach.

By building CDTs, we have been able to separate the impact of conduction velocity and heart anatomy on QRSd and quantitatively assess their relative changes across populations of different sex, BMI and age, which unveils mechanistic insights about the underlying biological process. Consistent with previous studies^35–38^, we found that men have longer QRSd and larger myocardial mass in comparison to women. This change in QRSd can be entirely explained by the change in anatomy, with no discernible disparity in CVs between the sexes, suggesting no significant biological differences in myocardium that modulate the ventricular depolarization wave propagation. Accordingly, clinical guidelines that use QRSd as criteria for elective therapy, such as cardiac resynchronization therapy, can develop sex-specific thresholds of decision based on heart size differences and disregard the potential impact of CV variations. This result brings additional evidence to support the proposal of reducing the threshold for CRT in female subjects by 9-13 ms^39^.

Similar to sex, longer QRSds are observed in overweight and obese groups compared to healthy groups, consistent with literature^37^, which are primarily attributed to the increased myocardial mass. Increased myocardial mass implies an increased length of the pathway for the ventricular activation wave to travel through, resulting in longer QRSd, which is consistent with previous studies showing obesity, as a common risk factor of cardiovascular diseases, can lead to structural remodelling, for example, left ventricular hypertrophy^40^. Obesity may also lead to electrical remodeling including conduction slowing and conduction heterogeneity, which is often observed in diseased clinical cohorts^41,42^. We have extended our knowledge of obesity-related electrical remodelling based on a relatively healthy cohort from the UK biobank. In contrast to earlier findings from clinical cohorts, we have observed a concurrent small but significant elevation in CVs corresponding to the increase in BMI. This suggests a potential adaptive response by the heart, possibly compensating for the effects of heightened myocardial mass on QRSd within obese groups. This novel insight, initially identified in our study, warrants further investigations to delve deeper into this adaptive mechanism and its implications.

We also observed longer QRSd in the elderly groups, which is attributed jointly to a decreased CV and an increased myocardial mass. This finding reaffirms previous research highlighting age-related changes including both structural changes such as increased LV wall mass^43^, possibly driven by cardiomyocyte hypertrophy and functional changes such as the decline in diastolic function^24,44^. This decline in the CV may be driven by well-established age-related cellular and tissue level remodelling including impaired sodium channel function^44^ (potentially though loss-of-function genetic mutations such as SCN5A as identified in previous ECG age-delta GWAS^45^ and experimental studies^46^) and the development of myocardial fibrosis^24,47^. It may also be affected by gap junction decoupling such as connexin 43 downregulation which is a known pathological remodelling in patients with ventricular hypertrophy and ischaemic heart diseases^48^.

PheWAS is a data-driven method to generate new hypotheses based on the identification of correlations between exposure like genetic and environmental factors and phenotypes like diseases and clinical outcomes. Unlike previous PheWAS studies focusing on either MRI-derived or genetic phenotypes^1,24,44^, our PheWAS study is the first to include an inferred tissue-level phenotype: CV, uniquely estimated with our CDT approach, which can form a bridge between research findings on the genetic, molecular/cellular level to tissue and the whole organ level.

Despite a relatively smaller sample size (N=3464), we found that biventricular myocardial mass was highly associated with multiple structural phenotypes such as LV stroke, end diastolic and end systolic volumes, as found in previously ^1^. Interestingly, we found that compared to the QRSd association with seven MRI-derived phenotypes, CV was only positively associated with LV ejection fraction, clinically used as a metric of functional performance particularly in the diagnosis of heart failure and post-myocardial infarction. Thus, CV may be used as a new biomarker for disease evaluation and also as a potential therapeutic target for guiding the development of new drugs.

Furthermore, we also identified notable correlations with multiple mental-health factors. The strongest correlations were observed between CV and QRSd with ‘the longest period of depression’ (QRSd: r=0.15 and CV:r=-0.15). Interestingly, this was not significantly associated with myocardial mass, although myocardial mass shows relatively weaker correlations with several other mental health factors (|*r*| < 0.1). Furthermore, in the separate association test on clinical outcomes, we found that reduced CV is more closely associated with an increased risk of both neurotic and mood disorders, in contrast to QRSd, while no significant results were observed for myocardial mass. Previous research has established a bidirectional relationship between depression and cardiovascular diseases, while a recent study specifically shows the association between low depression frequency with decreased risk of cardiometabolic disease using the UK Biobank data^49^. Increased evidence has linked psychological disorders with altered cardiac morphology and functions, for example, reduced LV mass, increased myocardial fibrosis and ^50,51^, which were also observed as changes in ECG metrics such as heart rate, QT interval, QRSd but not in all^52–54^. Our results reaffirmed these previous findings on myocardial mass and extend existing knowledge by identifying novel changes in CV and QRSd associated with neurotic and mood disorders (i.e. anxiety and depression etc).

Overall, the CDT-derived CVs exhibit greater sensitivity to functional alternations, compared to anatomical factors such as myocardial mass and ECG-derived metrics such as QRSd. This heightened sensitivity could be attributed to the fact that CV is directly modulated by the physical properties of cardiac myocytes and their interconnections. Therefore, the changes in CV may more accurately reflect cardiac remodelling such as fibrotic alterations resulting from the decoupling of cell-cell connections and coupling of myocytes with fibroblasts^55^.

In conclusion, we provide a proof-of-concept cross-sectional study that demonstrates the potential for applying a CDT workflow to larger cohorts. This approach may yield deeper insights into the biological connections between changes in cardiac morphology and function with neurological disorders. Further validation can be pursued through longitudinal studies utilizing resources such as the UK Biobank’s repeat imaging, and causal relationships can be established through large-scale genetic studies, such as the emerging area of heart-brain connection^56^.

## Methods

### Cardiac digital twin creation

The data used in this study is from the UK biobank, an open-access resource, under Application Number 88878. Ethical approval is obtained from the Northwest Research Ethics Committee (REC reference: 11/ NW/0382) and written consent is obtained from all participants. The full heart MRI protocol is described previously^1^. The present study was performed on the 4,329 first participants from the UK biobank who had adequate geometrical information^2^ and reported QRS duration (QRSd), sex, age, BMI/weight and height information. The details of the selection of population sample size and quality control were described previously^2,3^. The CDT creation workflow includes anatomical mesh construction and electrophysiological model calibration, and Supplementary Table 1 shows the summary participant characteristics for subjects who successfully went through both steps with existing QRSd, sex, age, BMI/weight, and height information reported in UKBB.

### Anatomical mesh generation

We used a nnU-net based architecture^4^ for automatic segmentation of the LV and RV blood pools and LV myocardium, trained based on manual segmentations on short-axis heart images^3^. The end-diastolic (ED) phase was selected as the first phase of acquisition. The contours and landmarks of LV and RV derived from the segmentations on the ED frame, were fed to an atlas-based pipeline (previously validated)^2^ to construct personalized biventricular surface meshes. The RV epicardium was estimated by extending the RV endocardium points normal to the surfaces by 3 mm consistent with experimental measurements^5,6^. The surface meshes were used to construct tetrahedral finite element meshes using Meshtool^7^ including regions of LV myocardium, RV myocardium, aortic, tricuspid, pulmonary and mitral valves. The biventricular myocardial mass was computed from the myocardial volume using a density of 1.05 *g*/*mL*.

To enable automated computation for CDTs, a morphological coordinate system, known as universal ventricular coordinates (UVCs), was introduced for describing positions within ventricles based on the apical-basal (*Z*), transmural (*ρ*) (from endocardium to epicardium), rotational (*φ*) (anterior, anteroseptal, inferior, inferolateral, anterolateral) and chamber-wise (left ventricle and right ventricle) coordinates. Biventricular myocardial fibre structure was implemented using a rule-based approach with a transmural variation of angle *α* as from 60° to -60° in longitudinal fibre directions and angle *β* as from −65° to 25° in transverse fibre directions from endocardium to epicardium.

### Electrophysiological (EP) model framework

The electrophysiological simulations are performed using the cardiac arrhythmia research package (CARP)^8^. We used a reaction-eikonal model without diffusion to compute the sinus ventricular activation times and transmembrane potential transient over time^9^. The activation wavefront propagation in the myocardium Ω is described as:

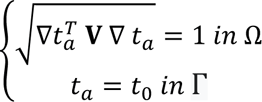

where *t*_*a*_ is the local activation time at any location in the myocardium, *t*_0_ are the instants of initial activation at locations Γ and the tense field **V** encodes the spatially heterogeneous orthotropic squared conduction velocity (CV). The ventricular myocardium was treated as transversely isotropic conductors. The ten Tusscher ionic model was used to simulate electrophysiological dynamics in the ventricular myocytes^10^. The ventricular depolarization during sinus rhythm is initiated by the His-Purkinje system (HPS). As direct measurement of the HPS in the cohort is not available, a fascicular-based model was used to represent the emergent physiological features of the HPS^11^. Overall, the electrophysiological framework consists of 20 EP parameters with uncertainty as shown in Extended Fig. 1 and Supplementary Table 2.

**Extended Fig. 1:**
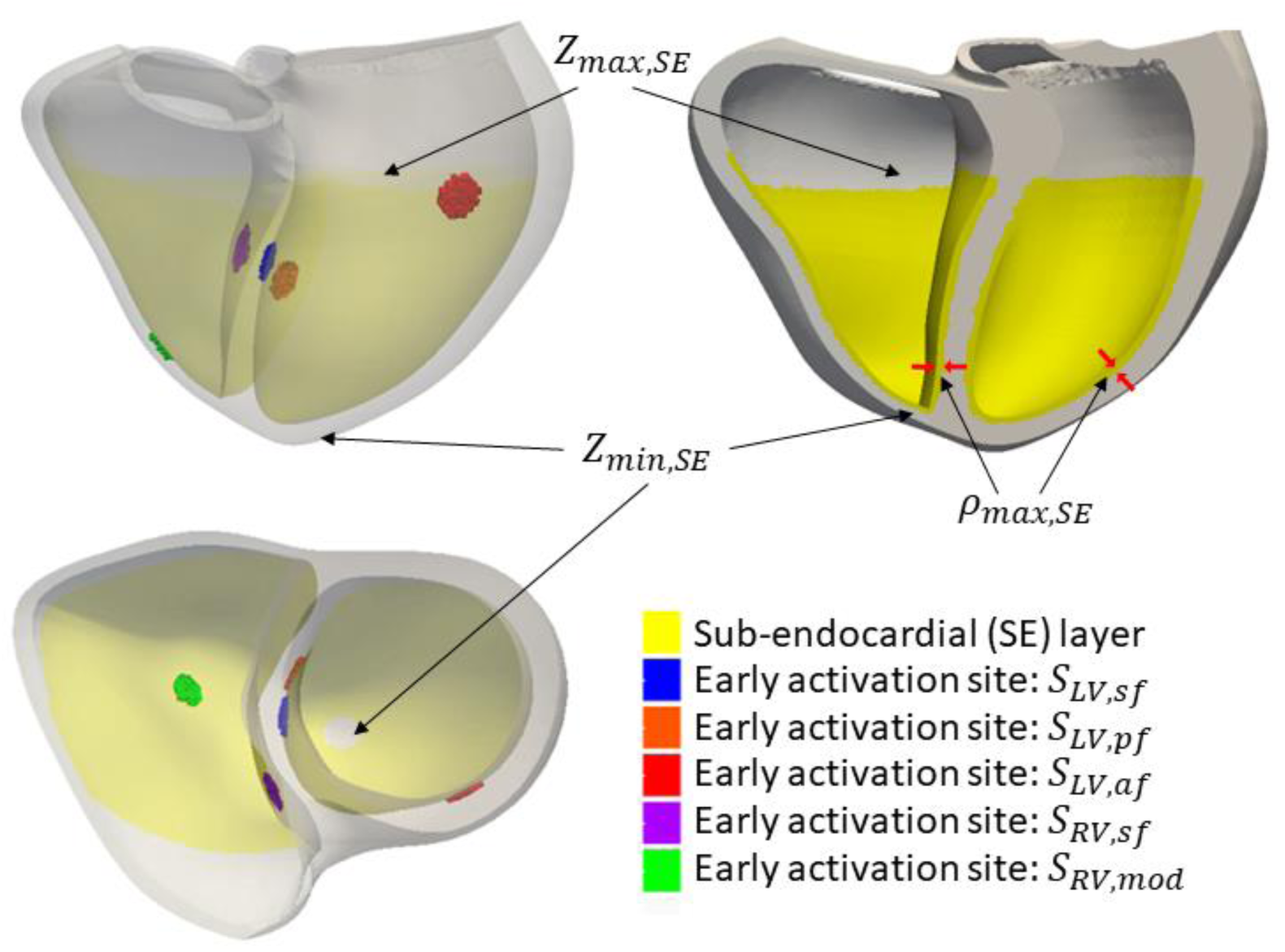
Fascicular-based model illustration replicating the realistic ventricular activation modulating by the His-Purkinje system. The SE layer (yellow) represents the fast conduction regions, where the fascicles are located, defined by apical-basal coordinate *Z* and transmural coordinate *ρ*, bounded by physiological measurements from literature as shown in Supplementary Table 2. There are also five early activation sites represented by disks with a thickness of 5% of the ventricular wall (**δ**_*z*_and **δ**_*ρ*_=0.05).) and having a fixed radius of 20*μm*, representing ∼25 cells. The disks are centred at root locations, defined by apical-basal coordinates *Z* and rotational coordinates *φ*, which are activated at specific timings. Both coordinates and timings of the five sites are bounded by physiological measurements from literature as shown in Supplementary Table 2.

Computing electrocardiograms (ECGs) requires information on the position of the heart within the torso, however, this information was not available. The heart models were therefore registered to a heart enclosed in an existing torso model^12^ using the UVCs. In this torso model, the locations of electrodes used in measuring 12-lead ECGs were identified the corresponding extracellular potentials were simulated, and the ECGs were computed. The QRSd was computed by finding the time points at which the spatial velocity exceeds 0.15 of the maximum spatial velocity in the reconstructed corresponding vectorcardiogram (VCG) from 12-lead ECGs, which fuse the information in all ECG traces^12^. This approach introduced extra uncertainty regarding the relative locations of the ten ECG electrodes within the real torsos. To quantify this uncertainty, we introduced another 30 parameters (Supplementary Table 3) which describe the variation of the cartesian coordinates for the ten electrodes and assumed that each cartesian coordinate can vary ± 5 *cm*, sufficiently to take account of all possible variations in electrode locations^13^.

In summary, the entire electrophysiological framework resulted in 50 parameters with uncertainty, including 20 tissue-level characteristics (Supplementary Table 2) and 30 additional parameters for the electrode locations (Supplementary Table 3).

### Gaussian process emulator and global sensitivity analysis

To build personalized CDTs, we need to set 50 parameters. However, the clinical measurements needed to constrain these parameters are not available and classical calibration techniques are prohibitive due to the massive computational costs required per case and the number of cases that we need to calibrate. We use Gaussian process emulators (GPEs) as surrogate models to accelerate the evaluation of the effect of the input parameters on the model output of interest: QRSd. This allows us to (1) gain important mechanistic knowledge about the input-output interactions and (2) through a GSA exclude the parameters that have little/no effects on the output, speeding up the personalization pipeline.

To investigate the effects of input parameters on QRSds in the cohort, we generated 10 representative samples by Latin hypercube sampling on basic characteristics including sex, age and BMI and identifying the 10 cases, who have the closest information to those samples, as representative subjects. Then we trained Gaussian processes emulators (GPEs) for each subject with the output as QRSd and performed a global sensitivity analysis (GSA) using the trained GPEs, to identify the key input parameters explaining the majority variation in the output QRSd. We train GPEs and perform GSA, firstly on the 20 tissue-level EP parameters to exclude parameters having small effects on the output QRSd. Then we combined the key input parameters with the other 30 parameters of ECG electrodes’ locations to identify the key input parameters in the whole EP framework, explaining the majority of variation in the output QRSd.

The training of GPEs is described previously^14^ by maximizing the model log-marginal likelihood using the GPErks emulation tool (http://github.com/stelong/GPErks). We evaluated the accuracy of GPEs using both coefficient of determination *R*^2^ and independent standard error *ISE* as:

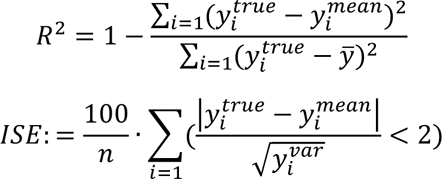

where *y_i_*^*true*^ is the true output, *ȳ* is the mean of true outputs, *y_i_*^*mean*^ and *y_i_*^*var*^ is the predicted posterior mean and variance of emulator outputs. The *R*^2^evaluates the error between the predictions and the observations with close to 1 indicating a lower error. The *ISE* accounts for the distance between predictions to the true values and quantifies the GPE uncertainty. If *ISE* is close to 100%, it means that the true values are falling in the region of the predictions within 2 standard deviations. The *R*^2^and *ISE* of trained GPEs for the 10 subjects were shown in Supplementary Table 4.

We then performed a variance-based global sensitivity analysis (GSA)^15^ on the trained GPEs to quantify the total effect for each input parameter (*S*_*T*_) on QRSd. The total effect consists of the first-order effect and the higher-order interactions, which are computed using the Saltelli method^15^ with the SALib Python library by taking N=1000 samples from the posterior distribution of trained GPEs. This allowed us to account for the effect of GPE uncertainty on the sensitivity indices. From the GSA, we ranked the total effects (*S*_*T*_) of the input parameters to identify the key parameters explaining the majority variation in the output QRSd. To train each GPE, we used Latin hypercube sampling to obtain 300 samples, and we ran EP simulations for those samples to build the training dataset. The impact of training data set size on the GSA was tested in the supplementary Figure 1.

### Personalised QRS duration calibration

To facilitate the replication of the clinically measured QRSd in the UK biobank across populations, a calibration workflow without any manual intervention is preferred. The GSA performed above enables us to identify the key input parameters responsible for the variation of QRSd, and allows us to reduce the number of input parameters required for the calibration process. Here we choose to only vary the most important parameter identified in the GSA for QRSd calibration, therefore allowing a computationally efficient bisection method to search for the subject-specific parameter (constrained by physiological measurements in literature) to match the corresponding QRSd.

To quantify the uncertainty brought by only varying the most important parameter to QRSd, we have computed confidence intervals for the inferred parameter for the 10 subjects used in GPE training. First, we randomly sampled (N=300) the other 49 parameters, assuming they are from a normal distribution with a mean equal to the median of the physiological bounds and the upper and lower bounds set at mean plus or minus three standard deviations (encompassing 99.72% of the data). Then, we performed simulations using the 300 samples combined with the subject-specific fitted parameter, to estimate the potential range of the output QRSd in each subject given the uncertainty in the uncalibrated 49 parameters. We then used the 5th and 95th percentiles of the output QRSd as the targets, to refit the key input parameter and then calculate their deviations from the original subject-specific fitted parameters which represents the confidence interval of the inferred parameter, considering the variation of other 49 parameters as in the Supplementary Figure 2.

### 12 lead ECGs comparison between the simulations and the recordings in UKBB

To assess the approximate errors of the simulated subject-specific 12 lead ECGs due to using a standard torso and fixed electrode locations in the personalized QRSd calibration, we compared the simulated 12 lead ECGs of 10 representative subjects across sex, age and BMI with their recorded ECGs in UKBB. The simulated 12 lead ECGs were first filtered by the same filters as used in processed recorded ECGs in the UKBB. It includes a low pass filter at 100 Hz, a high pass filter at 50 Hz and a notch filter at 50 Hz. Then, each lead of simulated ECGs was temporally aligned to the corresponding recorded lead ECG by matching the time points that the maximum energy was achieved (*V*^2^) and was also scaled in amplitude to obtain the same maximum absolute values as in the recorded ECG. Each lead of the recorded ECG was also cropped to have the same length as the aligned and scaled simulated lead ECG. Finally, each pair of simulated and recorded lead ECGs were compared by computing the Pearson correlation coefficient (*r*). For each subject, the 12-lead ECGs were ranked by *r* and the average *r* were calculated by considering different numbers of ranked ECG leads.

### Statistical analysis

Statistical analysis was performed using the Python Statsmodels library. The results were presented as mean ± a standard deviation unless specified. The body mass index (BMI) was calculated from height and weight measures taken at the time of the MRI being taken. Codes for the UK Biobank fields are included in brackets. Age was computed using the year of birth (34), month of birth (52) and date of attending the assessment centre (53) to get the actual age when imaging occurred. The Student t-test was used to compare two different groups. In the phenome-wide association (PheWAS) study, we used a similar approach as in^16^. Before computing univariate cross-correlation, effects such as age, sex, weight, and height were regressed out of the multimodal phenotypes, as they may confound with many phenotypes. The phenotypes from UKBB were normalized and then univariate cross-correlation was applied between the de-confounded multimodal phenotypes and the UKBB phenotypes. The UKBB phenotypes were categorized into 15 groups, including pulse wave analysis (128), LV size and function (133), abdominal composition (149), primary demographics (1001), early life (1002), self-reported medical conditions (1003), lifestyle diet (1004), physical measures (1006), education employment (1007), mental health (1018), summary diagnosis derived from summary diagnoses for hospital inpatient (41270), lifestyle alcohol (100051), physical activity (100054), smoking (100058), and medication. The self-reported medical condition and summary diagnosis were processed to have each column representing one disease code sorted in ascending order, before using in PheWAS. The medications group consists of all phenotypes relating to medications being used summarized from UKBB with codes including 20003, 22170, 22172, 22179, 22167, 22174, 22169, 22171, 22168, 22176, 22166, 22175, 22180, 22181, 22173, 22178, 22177, 6177, 6153, 10004, 6154, 20504, 20551, 20076, 20549, 20546, 2492. We cleaned the data before performing PheWAS by discarding phenotypes with more than 90% missing data and if two highly correlated phenotypes with correlation coefficient>0.9999, only one phenotype was kept.

## Data availability

The imaging data and non-imaging participant phenotypes and clinical outcomes are available from UK Biobank via a standard application procedure at http://www.ukbiobank.ac.uk/register-apply. The model generation are performed using open-sourced software: OpenCarp, available at https://opencarp.org/.

## Acknowledgements

This project is supported by the Wellcome Trust and EPSRC Centre for Medical Engineering (WT203148/Z/16/Z). S.N is also supported by NIH R01-HL152256, ERC PREDICT-HF 453 (864055), BHF (RG/20/4/34803), EPSRC (EP/X012603/1, EP/P01268X/1) and by the Technology Missions Fund under the EPSRC Grant EP/X03870X/1 & The Alan Turing Institute. M.B acknowledges BHF Project Grants PG/22/11159 and PG/22/10871.

## Author contributions

S.Q, M.S, R.R, A.Y, P.L, M.B, and S.N conceived and designed the study. S.Q, D.U, E.F, L.T, and Y.D implemented the anatomical model generation pipeline and applied it to the cohort. S.Q implemented the functional twinning pipeline, applied it to the cohort, analyzed the results and wrote the manuscript. All authors contributed to the manuscript revision and approved the final submitted version.

