## Supplementary material for "Developing Cardiac Digital Twins at Scale: Insights from Personalised Myocardial Conduction Velocity"

[ǀ] BioTechMed-Graz, Graz, Austria

[‖] University of Bordeaux, CNRS, Bordeaux, Talence, France

[ʃ] IHU Liryc, Bordeaux, Talence, France.

[§] Alan Turing Institute, British Library, 96 Euston Rd, London, UK

[‡] National Heart and Lung Institute, Imperial College London, London, UK

Corresponding author: Dr Qian, 4th North Wing, St Thomas' Hospital, London, SE1 7EH

### Abstract

Large-cohort studies using cardiovascular imaging and diagnostic datasets have assessed cardiac anatomy, function, and outcomes, but typically do not reveal underlying biological mechanisms. Cardiac digital twins (CDTs) provide personalized physics- and physiology- constrained *in-silico* representations, enabling inference of multi-scale properties tied to these mechanisms.

We constructed 3464 anatomically-accurate CDTs using cardiac magnetic resonance images from UK biobank and personalised their myocardial conduction velocities (CVs) from electrocardiograms (ECG), through an automated framework.

We found well-known sex-specific differences in QRS duration were fully explained by myocardial anatomy, as CV remained consistent across sexes. Conversely, significant associations of CV with ageing and increased BMI suggest myocardial tissue remodelling. Novel associations were observed with left ventricular ejection fraction and mental-health phenotypes, through a phenome-wide association study, and CV was also linked with adverse clinical outcomes.

Our study highlights the utility of population-based CDTs in assessing intersubject variability and uncovering strong links with mental health.

### Supplement

| | Total (n=3464) |
|---|---|
| Age (year) | 61.8 ± 7.5 |
| Male | 1600 |
| Female | 1864 |
| Caucasians | 3326 (95.6%) |
| BMI (kg/m$^2$) | 26.6 ± 4.4 |
| Diabetes | 164 (4.7%) |
| Hypertensive diseases | 807 (23.3%) |
| Ischaemic heart diseases | 283 (8.2%) |
| Heart failure | 60 (1.7%) |
| Fascicular block | 46 (1.3%) |
| Neurotic disorders (anxiety) | 139 (4.0%) |
| Mood [affective] disorder (depression) | 172 (5.0%) |

Supplementary Table 1: Basic characteristics of participants. N=3464. The body mass index (BMI) was calculated from height and weight measures taken at imaging. Age was computed using the year of birth (34), month of birth (52) and date of attending assessment centre (53) to get the actual age when imaging. Disease codes are derived from summary diagnoses in UKBB (41270) as shown in Supplementary Table 2. The percentages of specific subjects in the whole cohort are shown in brackets.

| Parameters | Range | Description | References |
|---|---|---|---|
| CV (m/s) | (0.48,0.92) | Myocardial conduction velocity along fiber | [1,2] |
| $Z_{LV,sf}$ | (0.25,0.75) | Apical-basal coordinate for LV septal fascicle | [3–6] |
| $Z_{LV,pf}$ | (0.25,0.75) | Apical-basal coordinate for LV posterior fascicle | [3–5] |
| $Z_{LV,af}$ | (0.6,0.8) | Apical-basal coordinate for LV anterior fascicle | [3–5] |
| $Z_{RV,sf}$ | (0.35,0.85) | Apical-basal coordinate for RV septal fascicle | [3–5] |
| $Z_{RV,mod}$ | (0.15,0.65) | Apical-basal coordinate for RV moderate band fascicle | [3–5] |
| $\varphi_{LV,sf}$ | (-1,1) | Rotational coordinate for LV septal fascicle | [3–6] |
| $\varphi_{LV,pf}$ | (-2,0) | Rotational coordinate for LV posterior fascicle | [3–6] |
| $\varphi_{LV,af}$ | (1.6,3.14) | Rotational coordinate for LV anterior fascicle | [3–6] |
| $\varphi_{RV,sf}$ | (0,2) | Rotational coordinate for RV septal fascicle | [3–6] |
| $\varphi_{RV,mod}$ | (-1,1) | Rotational coordinate for RV moderate band fascicle | [3–6] |
| $t_{LV,sf}$ | (0,30) | Firing time for LV posterior fascicle | [3] |
| $t_{LV,pf}$ | (0,30) | Firing time for LV anterior fascicle | [3] |

| | | | |
|---|---|---|---|
| $t_{LV,af}$ | (0,30) | Firing time for RV septal fascicle | 3 |
| $t_{RV,sf}$ | (0,30) | Firing time for RV moderate band fascicle | 3 |
| $t_{RV,mod}$ | (0,30) | Firing time for LV posterior fascicle | 3 |
| $\rho_{SE}$ | (0.01,0.3) | Transmural coordinate for SE layer | 7,8 |
| $Z_{min,SE}$ | (0.01,0.15) | The minimum apical-basal coordinate for SE layer | 6 |
| $Z_{max,SE}$ | (0.7,0.8) | The maximum apical-basal coordinate for SE layer | 6,9,10 |
| $R_{myo,SE}$ | (3,7) | The ratio of CV in SE layer to CV in myocardium | 7,8,11,12 |

Supplementary Table 2: 20 tissue-level electrophysiological characteristics with uncertainty with bounds based on literature. Note the myocardial CV transverse to fiber direction can also varied but is assumed to be proportion to the CV along the fiber in the table with the anisotropy ratio as 0.42 according to Ref[13]

| ECG Electrodes | Coordinate ranges |
|---|---|
| RA | $X_{RA,recovered} \pm 5\ cm, Y_{RA,recovered} \pm 5\ cm, Z_{RA,recovered} \pm 5\ cm$ |
| LA | $X_{LA,recovered} \pm 5\ cm, Y_{LA,recovered} \pm 5\ cm, Z_{LA,recovered} \pm 5\ cm$ |
| LL | $X_{LL,recovered} \pm 5\ cm, Y_{LL,recovered} \pm 5\ cm, Z_{LL,recovered} \pm 5\ cm$ |
| RL | $X_{RL,recovered} \pm 5\ cm, Y_{RL,recovered} \pm 5\ cm, Z_{RL,recovered} \pm 5\ cm$ |
| V1 | $X_{V1,recovered} \pm 5\ cm, Y_{V1,recovered} \pm 5\ cm, Z_{V1,recovered} \pm 5\ cm$ |
| V2 | $X_{V2,recovered} \pm 5\ cm, Y_{V2,recovered} \pm 5\ cm, Z_{V2,recovered} \pm 5\ cm$ |
| V3 | $X_{V3,recovered} \pm 5\ cm, Y_{V3,recovered} \pm 5\ cm, Z_{V3,recovered} \pm 5\ cm$ |
| V4 | $X_{V4,recovered} \pm 5\ cm, Y_{V4,recovered} \pm 5\ cm, Z_{V4,recovered} \pm 5\ cm$ |
| V5 | $X_{V5,recovered} \pm 5\ cm, Y_{V5,recovered} \pm 5\ cm, Z_{V5,recovered} \pm 5\ cm$ |
| V6 | $X_{V6,recovered} \pm 5\ cm, Y_{V6,recovered} \pm 5\ cm, Z_{V6,recovered} \pm 5\ cm$ |

Supplementary Table 3: 30 ECG electrode positions characteristics with uncertainty.

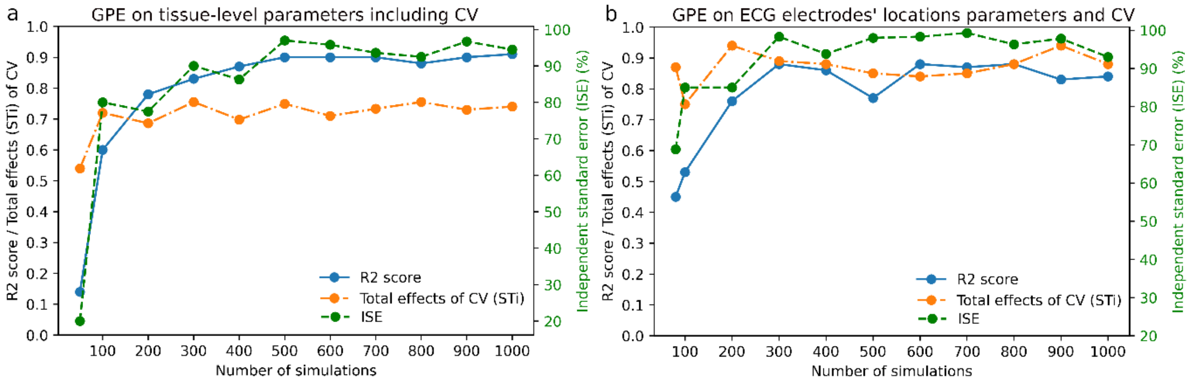


Supplementary Figure 1: Trained GPE validation

| | GPE on tissue-level parameters | | GPE on electrodes' locations | |
|---|---|---|---|---|
| | $R^2$ score | ISE (%) | $R^2$ score | ISE (%) |
| Subject 1 | 0.87 | 90 | 0.66 | 95.1 |
| Subject 2 | 0.78 | 90 | 0.41 | 98.3 |
| Subject 3 | 0.76 | 91.7 | 0.45 | 93.4 |
| Subject 4 | 0.83 | 90.7 | 0.51 | 97.7 |
| Subject 5 | 0.8 | 83.6 | 0.49 | 88.5 |
| Subject 6 | 0.79 | 90 | 0.56 | 90 |
| Subject 7 | 0.78 | 91.7 | 0.65 | 96.7 |
| Subject 8 | 0.83 | 88.3 | 0.74 | 91.7 |
| Subject 9 | 0.78 | 83.3 | 0.74 | 98.3 |
| Subject 10 | 0.91 | 94.5 | 0.88 | 98.3 |

| | | | | |
|---|---|---|---|---|
| Average $\pm Std$ | $0.81 \pm 0.04$ | $89.4 \pm 3.3$ | $0.63 \pm 0.15$ | $94.3 \pm 3.5$ |

Supplementary Table 4: $R^2$ $score$ and independent standard error (ISE) for GPEs trained on 10 subjects, first on tissue-level parameters then on ECG electrodes' locations and the key parameters identified affecting the QRS duration most in the first GPEs.

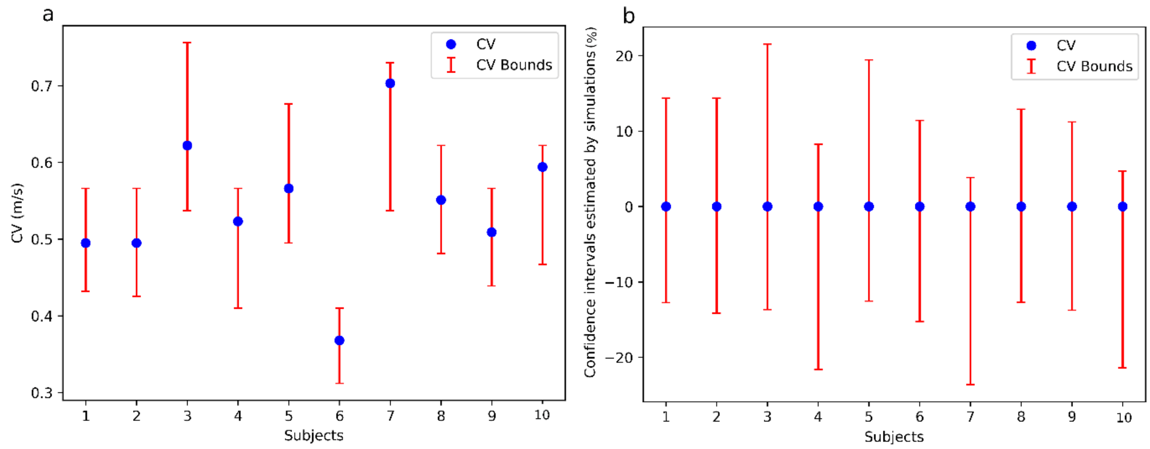


Supplementary Figure 2: a. The fitted CVs with error bars computed, considering the effect of uncertainty of the other 49 parameters on QRS duration, for the 10 subjects used in GPE. B. the corresponding confidence interval of fitted CVs in %.

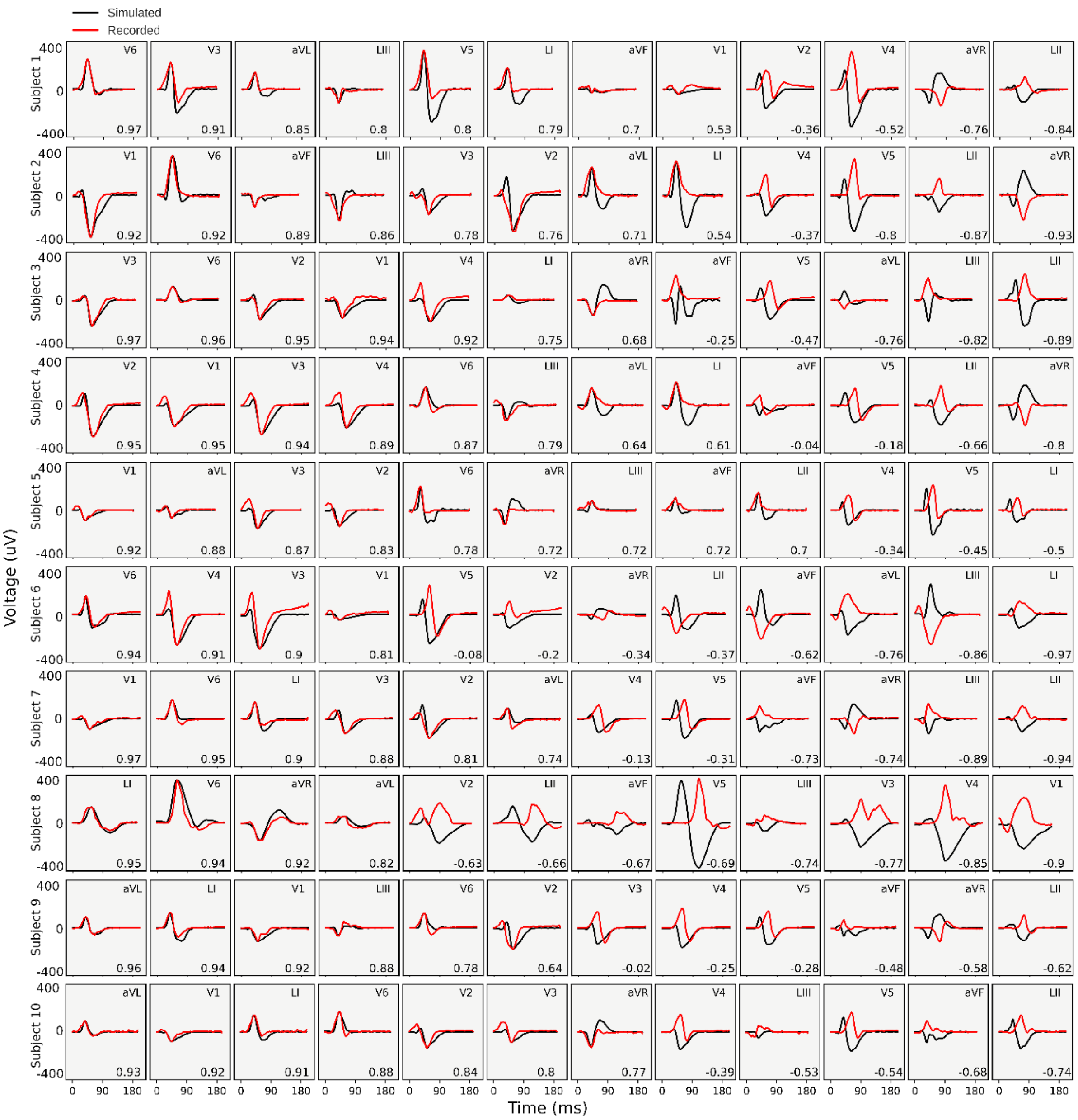


Supplementary Figure 3: The simulated 12 lead ECGs (black) of the ten sample subjects (same as in Figure 2) are compared with recorded ECGs in the UKBB (red). Each simulated lead ECGs were scaled to have the same maximum absolute amplitude as the corresponding recorded lead ECGs in $\mu V$. All lead simulated and recorded ECGs were temporally aligned by matching the timepoints that the maximum voltage energy were achieved. Then the correlation scores for each pair of simulated and recorded lead ECGs were computed as shown at the bottom right corner of the plots and the 12 lead ECGs were listed in descending order of their correlation scores.

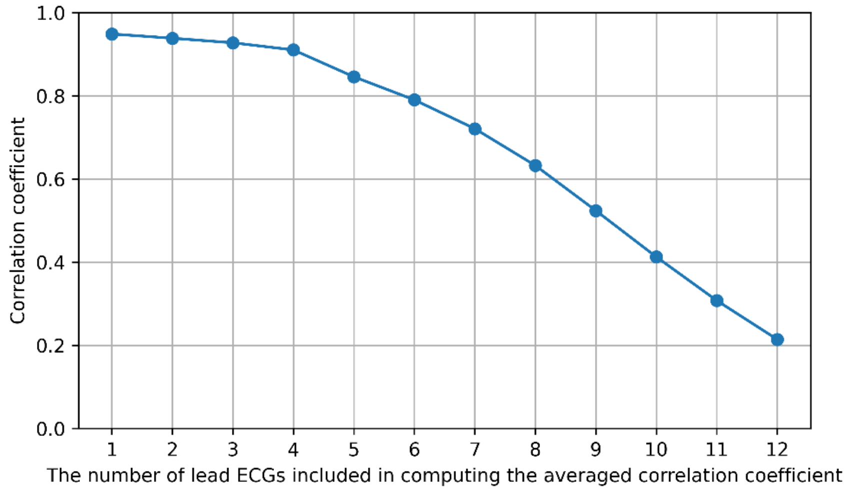


Supplementary Figure 4: The averaged correlation coefficients ($r$) of simulated and recorded ECGs versus the number of lead ECGs included in computing the averaged $r$.

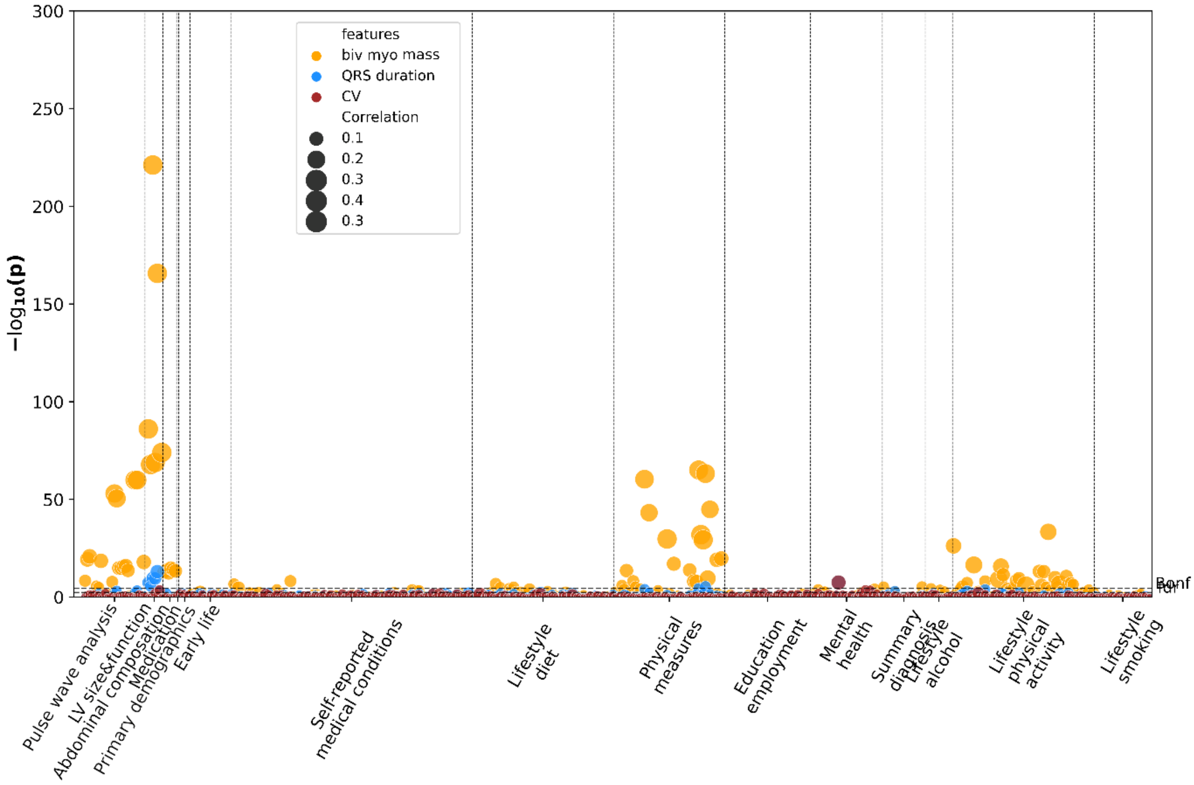


Supplementary Figure 5: Manhattan plot showing the $-log_{10}P$ (two-sided t-test) for correlations between the multimodal phenotypes and phenotypes in UKBB. The size of the dots indicates the absolute Pearson's correlation coefficient. The dashed horizontal lines are the Bonferroni threshold (Bonf) and the false-discovery rate (fdr) (α = 0.05).

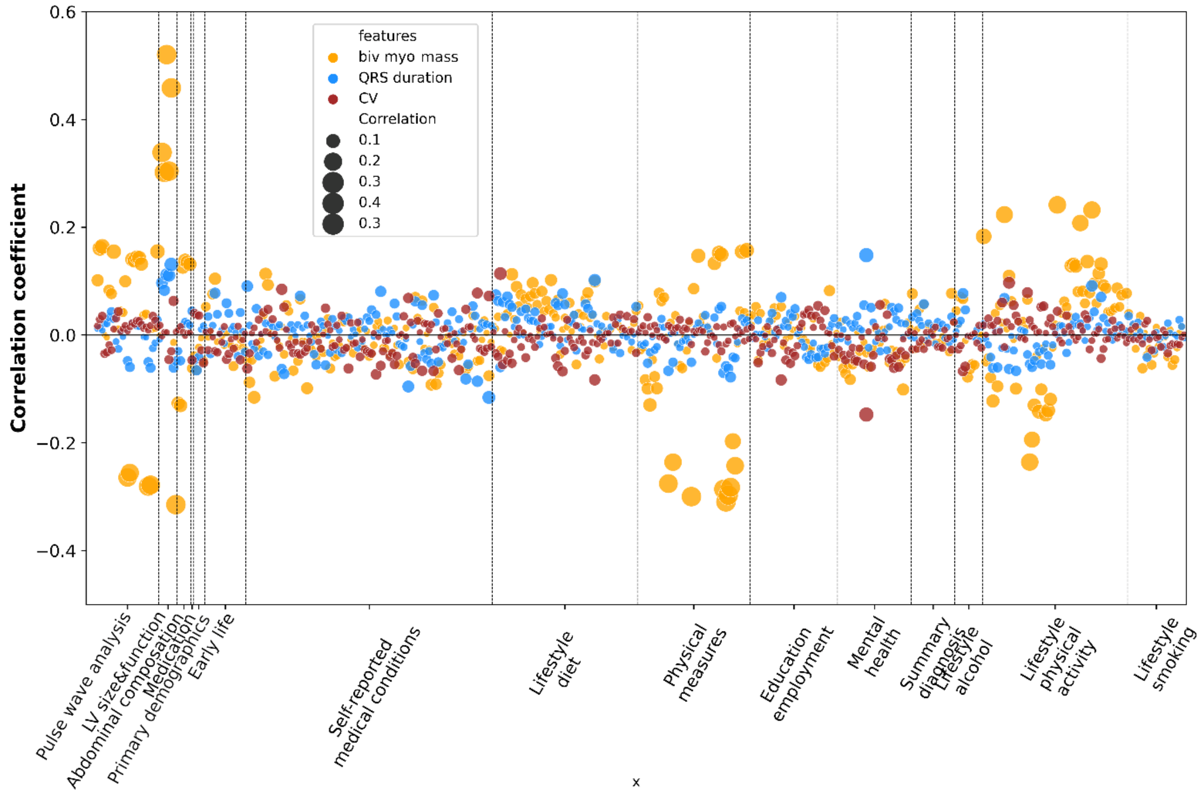


Supplementary Figure 6: The corresponding Manhattan plot as Sup figure 4 shows the correlation coefficients between the multimodal phenotypes and phenotypes in UKBB. The size of the dots indicates the absolute Pearson's correlation coefficient.

| Disease type | Meaning |
|---|---|
| Heart failure | I50.0 Congestive heart failure<br>I50.1 Left ventricular failure<br>I50.9 Heart failure, unspecified |
| Fascicular block | I44.4 Left anterior fascicular block<br>I44.7 Left bundle-branch block, unspecified<br>I44.5 Left posterior fascicular block<br>I45.0 Right fascicular block<br>I44.6 Other and unspecified fascicular block<br>I45.1 Other and unspecified right bundle-branch block<br>I45.2 Bifascicular block<br>I45.3 Trifascicular block<br>I45.4 Nonspecific intraventricular block |
| Ischaemic heart disease | I20-I25 Ischaemic heart diseases<br>I20 Angina pectoris<br>I21 Acute myocardial infarction<br>I22 Subsequent myocardial infarction<br>I23 Certain current complications following acute myocardial infarction<br>I24 Other acute ischaemic heart diseases<br>I25 Chronic ischaemic heart disease |
| Diabetes | E10-E14 Diabetes mellitus<br>E10 Insulin-dependent diabetes mellitus<br>E11 Non-insulin-dependent diabetes mellitus |

| | |
|---|---|
| | E12 Malnutrition-related diabetes mellitus<br>E13 Other specified diabetes mellitus<br>E14 Unspecified diabetes mellitus |
| Neurotic disorders | F40-F48 Neurotic, stress-related and somatoform disorders<br>F40 Phobic anxiety disorders<br>F41 Other anxiety disorders<br>F42 Obsessive-compulsive disorder<br>F43 Reaction to severe stress, and adjustment disorders<br>F44 Dissociative [conversion] disorders<br>F45 Somatoform disorders<br>F48 Other neurotic disorders |
| Mood disorders | F30-F39 Mood [affective] disorders<br>F30 Manic episode<br>F31 Bipolar affective disorder<br>F32 Depressive episode<br>F33 Recurrent depressive disorder<br>F34 Persistent mood [affective] disorders<br>F38 Other mood [affective] disorders<br>F39 Unspecified mood [affective] disorder |
| Hypertension | I10-I15 Hypertensive diseases-<br>I10 Essential (primary) hypertension<br>I11 Hypertensive heart disease<br>I12 Hypertensive renal disease<br>I13 Hypertensive heart and renal disease<br>I15 Secondary hypertension |

Supplementary Table 5: Common diseases categorized from summary diagnoses in UKBB (41270).

| | Heart failure | Fascicular block | Ischeamic disease | Diabetes | Neurotic disorder | Mood disorder | Hypertension |
|---|---|---|---|---|---|---|---|
| CV | 1.22$_{[1.05,1.4]}$ | 0.43$_{[0.37,0.51]}$ | 1.21$_{[1.06,1.37]}$ | 0.75$_{[0.66,0.86]}$ | 0.65$_{[0.57,0.73]}$ | 0.62$_{[0.54,0.72]}$ | 0.82$_{[0.71,0.94]}$ |
| QRS durtaion | 1.38$_{[1.20,1.58]}$ | 1.13$_{[0.98,1.29]}$ | 1.34$_{[1.19,1.52]}$ | 0.82$_{[0.71,0.94]}$ | 0.77$_{[0.68,0.88]}$ | 0.76$_{[0.66,0.87]}$ | 0.87$_{[0.75,1.01]}$ |
| Myo vol | 1.66$_{[1.50,1.83]}$ | 1.72$_{[1.54,1.92]}$ | 0.98$_{[0.88,1.08]}$ | 0.85$_{[0.76,0.95]}$ | 0.89$_{[0.8,0.99]}$ | 0.88$_{[0.79,0.98]}$ | 1.33$_{[1.18,1.49]}$ |
| Sex | 4.35$_{[2.18,8.68]}$ | 6.03$_{[3.2,11.36]}$ | 1.13$_{[0.68,1.88]}$ | 1.55$_{[0.93,2.58]}$ | 0.31$_{[0.19,0.48]}$ | 0.48$_{[0.3,0.75]}$ | 1.32$_{[0.75,2.32]}$ |
| Age | 1.77$_{[1.10,2.85]}$ | 0.36$_{[0.22,0.59]}$ | 2.25$_{[1.56,3.25]}$ | 0.85$_{[0.57,1.26]}$ | 1.29$_{[0.95,1.75]}$ | 0.53$_{[0.38,0.74]}$ | 2.73$_{[1.79,4.17]}$ |
| BMI | 0.43$_{[0.22,0.81]}$ | 0.04$_{[0.02,0.09]}$ | 1.66$_{[1.02,2.68]}$ | 1.13$_{[0.68,1.87]}$ | 2.16$_{[1.44,3.23]}$ | 1.10$_{[0.71,1.70]}$ | 2.14$_{[1.24,3.69]}$ |
| Age * BMI | 3.86$_{[1.82,8.22]}$ | 41.81$_{[18,97.1]}$ | 0.78$_{[0.43,1.39]}$ | 2.75$_{[1.49,5.08]}$ | 0.57$_{[0.35,0.95]}$ | 1.72$_{[1.00,2.96]}$ | 0.73$_{[0.37,1.41]}$ |
| Sex * Age | 0.25$_{[0.13,0.48]}$ | 0.14$_{[0.07,0.25]}$ | 1.14$_{[0.69,1.87]}$ | 0.92$_{[0.56,1.52]}$ | 3.09$_{[1.98,4.82]}$ | 1.82$_{[1.15,2.88]}$ | 0.84$_{[0.48,1.46]}$ |

Supplementary Table 6: Odd ratios [95% confidence interval] for phenotypes as a risk factor for a common disease as the outcome. The logistic regression analysis is adjusted for Sex, age, BMI, Age*BMI and Sex*Age as additional independent variables.

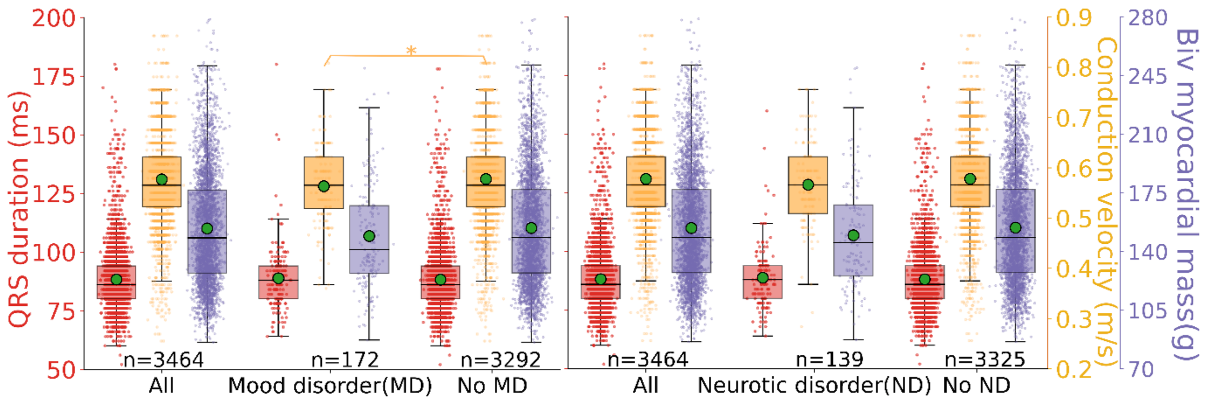


Supplementary Figure 7: Boxplots of QRS duration (red), conduction velocity (yellow) and myocardial mass (purple) for comparing groups of participants afflicted with mood disorder and neurotic disorder and non-afflicted counterparts. The green dots indicate the mean. The corresponding P values are from student's t-test. *P<0.05; **P<0.01; ***P<0.001; ****P<0.0001.
